# The utility of quantitative D-Dimer assay as a biomarker in the diagnosis and exclusion of Cerebral Venous Sinus Thrombosis

**DOI:** 10.1101/2022.12.21.22283814

**Authors:** Dipesh Soni, Ashok Kumar Pannu, Atul Saroch, Vikas Bhatia, Jasmina Ahluwalia, Rajveer Singh, Arihant Jain

## Abstract

Thrombotic disorders are characterized by the presence of elevated levels of detectable fibrin degradation products in the blood. The utility and sensitivity of quantitative D-Dimer assay to rule out the diagnosis of deep vein thrombosis is well established. We extrapolated this principle to evaluate the utility of D-Dimer assay in exclusion of cerebral venous sinus thrombosis (CVST). CVST is an important cause of cerebrovascular accidents in young patients and the residual neurological deficits can be minimized if correct therapy, i.e., anticoagulation is instituted in a timely manner. As advanced imaging modalities that are required for the diagnosis of CVST might not be readily available everywhere, it is important to have a sensitive biomarker which can guide clinicians to rule out the diagnosis with a reasonable confidence. We evaluated the patients admitted at a tertiary care center who underwent Computed tomography (CT) Venography/Magnetic resonance (MR) Venography of the brain with the clinical suspicion of CVST. After appropriate exclusion, a quantitative D-Dimer assay was performed in patients who had CVST on CT/MR Venography and was compared with those patients who did not. Receiver operating characteristic (ROC) analysis revealed that quantitative D Dimer had poor diagnostic accuracy for the differentiation of CVST from non CVST cases (Area under the curve = 0.694), but D-Dimer levels of <300 ng/mL had a sensitivity of 90% for ruling out the diagnosis of CVST.

## Introduction

Cerebral venous sinus thrombosis is understood as any thrombotic event taking place in the intracerebral veins or sinuses. It is an uncommon cause of stroke with nonspecific clinical presentation, various risk factors, different radiological findings and varied outcomes. Prompt recognition, early diagnosis and initiation of anticoagulation therapy can significantly reduce the incidence of acute and long-term complications. [1] Lack of a proper dedicated CVST registry bereft us of exact incidence & prevalence of CVST, but with the introduction of sophisticated radiological techniques, the incidence of CVST has increased in the past few years. Children, younger adults and females of reproductive age group from low-income countries are the most commonly affected population group. [2] There are no population-based studies in India, and with limited evidence from hospital based studies, a varied incidence of CVST seems apparent. Hospital-based series from Northern India in the late 1970s reported CVST in 4.5/1000 obstetric admissions. [3] In an autopsy series by Banerjee et al., CVST accounted for almost 10% of all strokes reported in their study. [4] A closer look at the available data from different case series suggests that the prevalence of CVST in India is comparable, if not more, than the reported prevalence in the western countries. The mechanism of neuronal injury in CVST differs significantly from that of the arterial occlusive diseases. In arterial occlusion, the nutrient and oxygen supply directly get cut off which leads to subsequent cerebral ischemia and infarction. When the venous system is occluded, drainage of blood is compromised. Hydrostatic pressure increases in vessels that were being drained by the now obstructed veins and sinuses, and cerebral edema develops in the involved territory. If the pressure is not relieved promptly, cerebral haemorrhage may ensue owing to capillary fragility at elevated pressures. The significant radiological findings that are present in patients of CVST include localized cerebral edema and areas of hemorrhage. The bleed can also spill out into the nearby subarachnoid space. [5]

The venous drainage of brain is taken care of by two systems, the superficial and the deep venous systems. The superficial drainage system is highly collateralized and drained into the Superior sagittal sinus and the Lateral sinus and the diagnosis of occlusion in this drainage system solely on clinical grounds is an arduous task. The deep venous system is responsible for draining the basal ganglia, deep white matter tracts towards the vein of Galen. Many anastomoses have been identified between the two drainage systems; however, the presence of anastomoses is not consistent and may be absent or poorly developed. (6) Prior to the era of antibiotics, most cases of CVST were due to infections involving the meninges, paranasal sinuses, face, ear, head and neck. In today’s world, where the threshold for initiating an antibiotic for a suspected infection is low, infections no more stand on top of the list as a potential causes of CVST. A person’s genetic susceptibility and environmental triggers push the individual towards developing the procoagulant state and thus, increases the chances of the individual having a thrombotic phenomenon.[7]

Before the advent of sophisticated imaging techniques such as computed tomography(CT) and magnetic resonance imaging (MRI), CVST was considered by many as a fatal disease that usually was diagnosed at autopsy. although this problem was significantly overcome by the introduction of advanced imaging modalities, widespread access and logistics remain a problem. Given the fact that CVST has a non-specific clinical presentation and examination findings, evaluating patients at a large volume centre can be challenging. Hence there is a need for a bedside point of care testing/diagnostic scoring for identification and timely management of CVST. [6]

The treatment strategy for CVST aims to reduce the symptoms of raised intracranial pressure secondary to venostasis, along with the management of seizures and the deficits that may come into the picture with development of cerebral edema and infarction. Anticoagulation is the cornerstone of treatment and is the standard of practice unless any contraindication to its use is present. Surgical and endovascular techniques are employed in situations where anticoagulants are ineffective. [7] Long term complications observed at follow up of these patients include extensive venous collateral formation, Dural AV fistula formation, raised ICP, which can cause herniation and permanent neurological deficits such as blindness or incoordination. [8] This study aims to evaluate the utility of quantitative D Dimer assay in the diagnosis and exclusion of CVST and to devise a clinical decision rule and for the diagnosis if CVST.

## Methods

### Study design

This is a hospital based, prospective, observational study that was carried out at tertiary care centre in northern India. The study was carried out for a period of 17 months between June 2020 to December 2021.

#### Inclusion criteria

1. Adult patients (age > 18 years) with suspected cerebral venous thrombosis Undergoing CT Venography (CTV) or angiography (CTA)/Magnetic resonance Venography (MRV) or angiography(MRA).
2. Patient with symptom duration less than seven days.

#### Exclusion criteria

1. Patients not giving consent for evaluation
2. Patients with symptom duration more than seven days.
3. Patients with proven/known thromboembolic disease (e.g., Deep vein thrombosis, pulmonary embolism) within the past four weeks.
4. Patients with Disseminated intravascular coagulation

Patients presenting with clinical features that raise the suspicion for cerebral venous sinus thrombosis, namely, 1. headache, 2. focal neurological deficits in the form of hearing disturbances, visual disturbances, motor and sensory deficits, 3. altered sensorium, 4. Seizures were planned for further diagnostic evaluation using CTV/CTA or MRV/MRA. At the end of the aforementioned investigation, the patients were divided into two groups, those that had features of CVST on radiological imaging (positive cases) and those who did not have any imaging findings suggestive of CVST (negative cases). A blood sample of 2.7 ml in a 3.2% sodium citrate vial was collected for quantitative D dimer levels. The blood sample was drawn before the patient injected with IV contrast, thus, any possible interference that may be because of the contrast was avoided. D Dimer estimation was done using the commercially available HemosIL D Dimer Hs kit, which is an enhanced latex immunoassay, and has a sensitivity of 95% and a specificity of 53% on the ACL TOP 500 CTS Coagulation analyser (Instrumentation Lab, USA). The turnaround time was 15 minutes, and the method is automated, with the only disadvantage being moderate specificity. [9] Patients who had CVST were subjected to further workup for the aetiology of CVST and were counselled regarding the nature of their disease.

### Statistical analysis

Demographic information was collected in a predetermined checklist. Sample size calculation was done using the Buderer formula (10) and a total of 83 participants were enrolled into the study. Categorical data was represented as n (%), Continuous data was presented as its mean and standard deviation or in the form of its median and interquartile range as per the normality of data. Chi-square/Fischer’s test was applied for comparison of the categorical variables as appropriate. Binary logistic regression was used to test the significance between D Dimer values and Positive/negative Cases of CVST. ROC analysis and area under the curve (AUC) was computed to determine the optimum cut-off value for exclusion of CVST. All the comparisons were at a 5% significance level, i.e., p <0.05, was considered significant. Data was electronically entered using a digital spreadsheet by MICROSOFT EXCEL (version 16.49). Analysis of data was performed by using IBM SPSS STATISTICS (version 22.0)

## Results

83 participants were enrolled in the study, of which most were females (n=47, 56%). 49 participants were diagnosed with CVST (positive cases) while 34 did not have any demonstrable thrombus on radiology (negative cases). Female participants constituted the higher proportion of CVST positive cases (65.3%, P=0.055).

A significantly larger number of patients with CVST complained of headaches as compared to those without CVST. (83.6% vs 52.9%, P=0.002). The presence of visual disturbances (24.5% vs 17.6%), seizures (55 % vs 50 %) and altered mental status (70% vs 20%) was also more in patients with CVST as compared to those without CVST, however, the difference between the two groups was not statistically significant. Among 49 patients of CVST, 24(48.9%) patients had no identifiable risk factor for CVST. Out of the remaining 25 participants, postpartum state (16%), hyperhomocysteinemia (10%) and presence of a connective tissue disease (6%) were common identifiable risk factors.

Patients with CVST had a higher level of quantitative D Dimer than those in the non CVST group, and this difference was statistically significant. (876.27 ng/mL, IQR 486-1642 vs 538.5 ng/mL, IQR 327-775, P=0.003). The Box-and-Whisker plot (Fig 1) depicts the distribution of Quantitative D-dimer (ng/mL). The middle horizontal line represents the median Quantitative D-dimer (ng/mL), the upper and lower bounds of the box represent the 75th and the 25th centile of Quantitative D-dimer (ng/mL), respectively, and the upper and lower extent of the whiskers represent the range of Quantitative D-dimer (ng/mL) in each of the groups.

**Fig 1.**
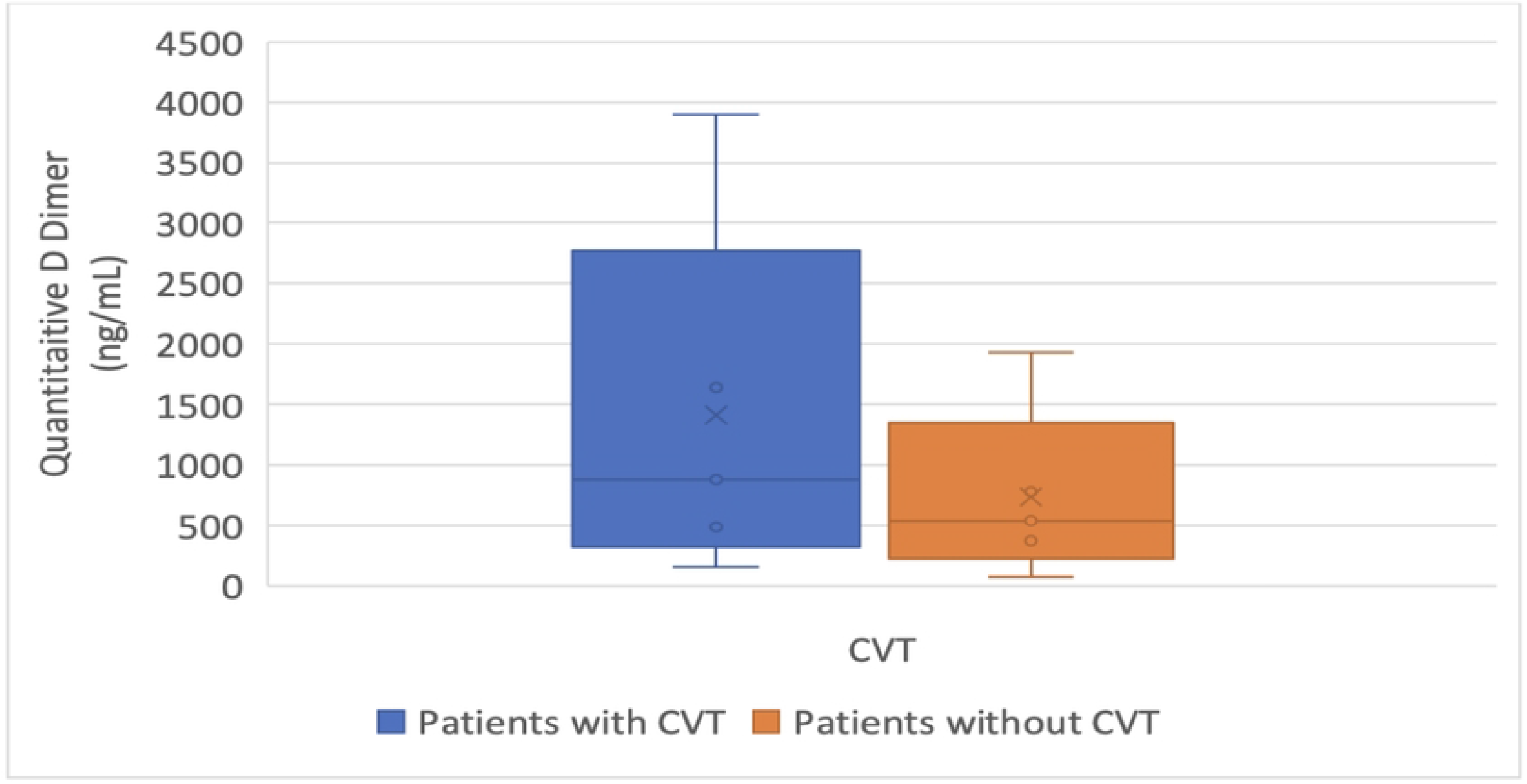
Distribution of Quantitative D Dimer levels in study participants.

ROC analysis was carried out to determine the diagnostic performance of Quantitative D-Dimer assay in Predicting CVST. With an AUROC of 0.694, quantitative D-Dimer Assay has a poor diagnostic performance in differentiating CVST from non-CVST patients. At a D-Dimer value of ≥792 ng/mL, the diagnosis of CVST was likely with a specificity of 79% and a sensitivity of 57%, however, at a value of <300 ng/mL, D-Dimer had a sensitivity of 89.8% for the exclusion of the diagnosis of CVST. (Fig 2), (Table 1)

**Table 1:** ROC curve analysis showing diagnostic performance of quantitative D-dimer (ng/mL) in predicting CVST (n = 83)

| Parameter | Value (95% CI) |
| --- | --- |
| Cutoff (p value) | $\geq 792$ (0.003) |
| AUROC | 0.694 (0.58 - 0.809) |
| Sensitivity | 57.1% (42-71) |
| Specificity | 79.4% (62-91) |
| Positive Predictive Value | 80.0% (63-92) |
| Negative Predictive Value | 56.2% (41-71) |
| Diagnostic Accuracy | 66.3% (55-76) |
| Positive Likelihood Ratio | 2.78 (1.37-5.61) |
| Negative Likelihood Ratio | 0.54 (0.37-0.78) |
| Diagnostic Odds Ratio | 5.14 (1.88-14.06) |

**Fig 2:**
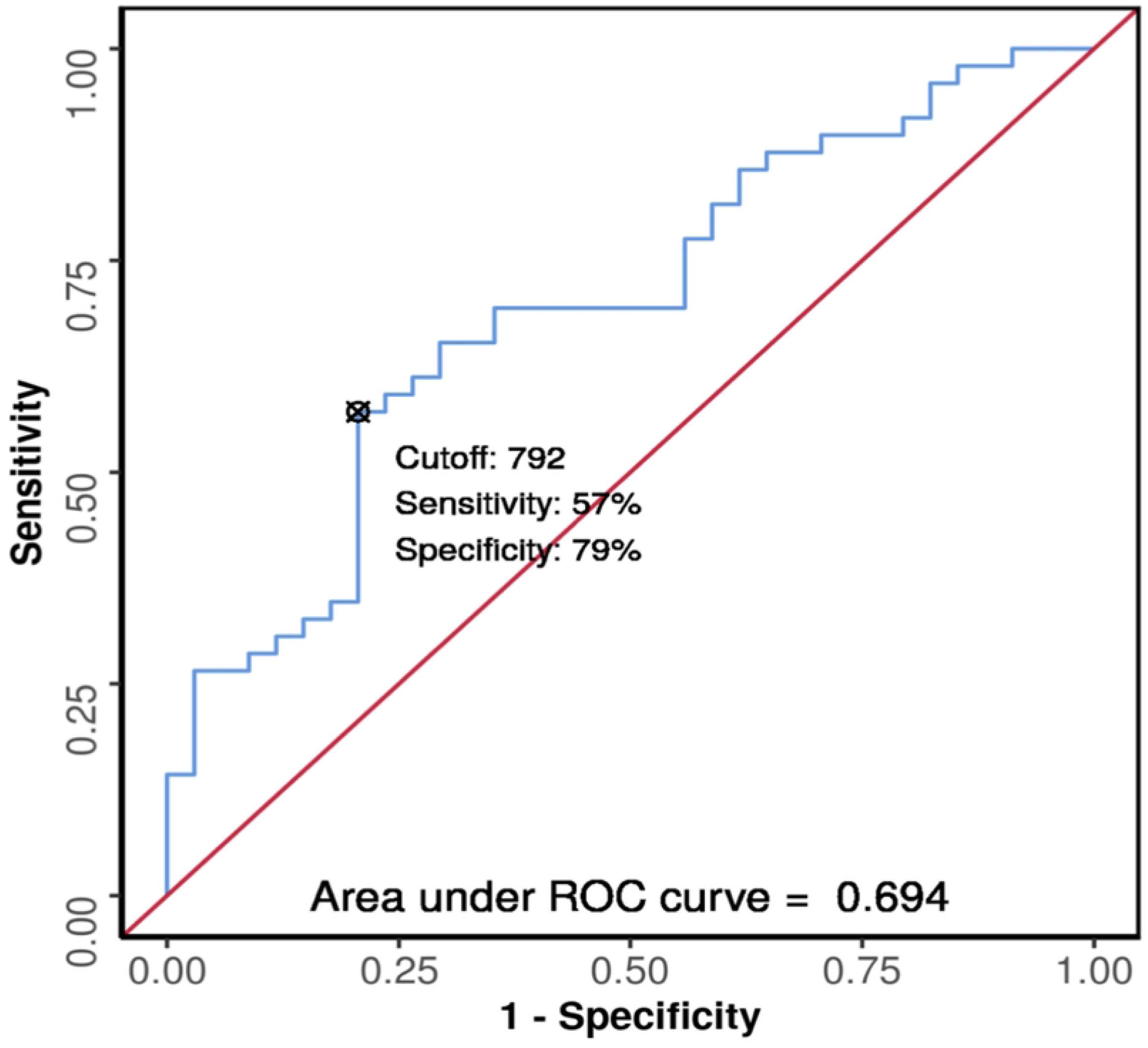
ROC curve analysis showing diagnostic performance of quantitative D-dimer (ng/mL) in predicting CVST (n = 83)

To devise a clinical decision rule, patient factors which had a strongest association with CVST were identified. After univariate analysis of the data, presence of headache and a D-Dimer level of ≥792 ng/mL were significantly associated with the presence of CVST. Other than the above two, Female gender was also included in the computation of the decision rule as most of the studies from the recent past have demonstrated that female gender confers a higher relative risk for the development of CVST. [1, 11, 12, 13].

A regression analysis for the diagnosis of CVST was conducted, with the dependent variable being CVST using all the above mentioned three factors. Multivariate logistic regression was carried out to identify the odds ratio for all the factors. Following table (Table 2) summarizes the regression analysis for the dependent variable, CVST, using all the predictor variables together in one go. The ‘OR (univariable)’ column lists the odds ratios for each of the variables with respect to the dependent variable, when these variables are used as single predictors of the dependent variable, without entering the rest of the variables in the model. The ‘OR (multivariable)’ column lists the odds ratios for all the variables when they are entered in the model together (and are now thus controlling for each other). The first category in each of the categorical variables is the reference category, against which the odds ratios of the rest of the variables is calculated.

**Table 2:**
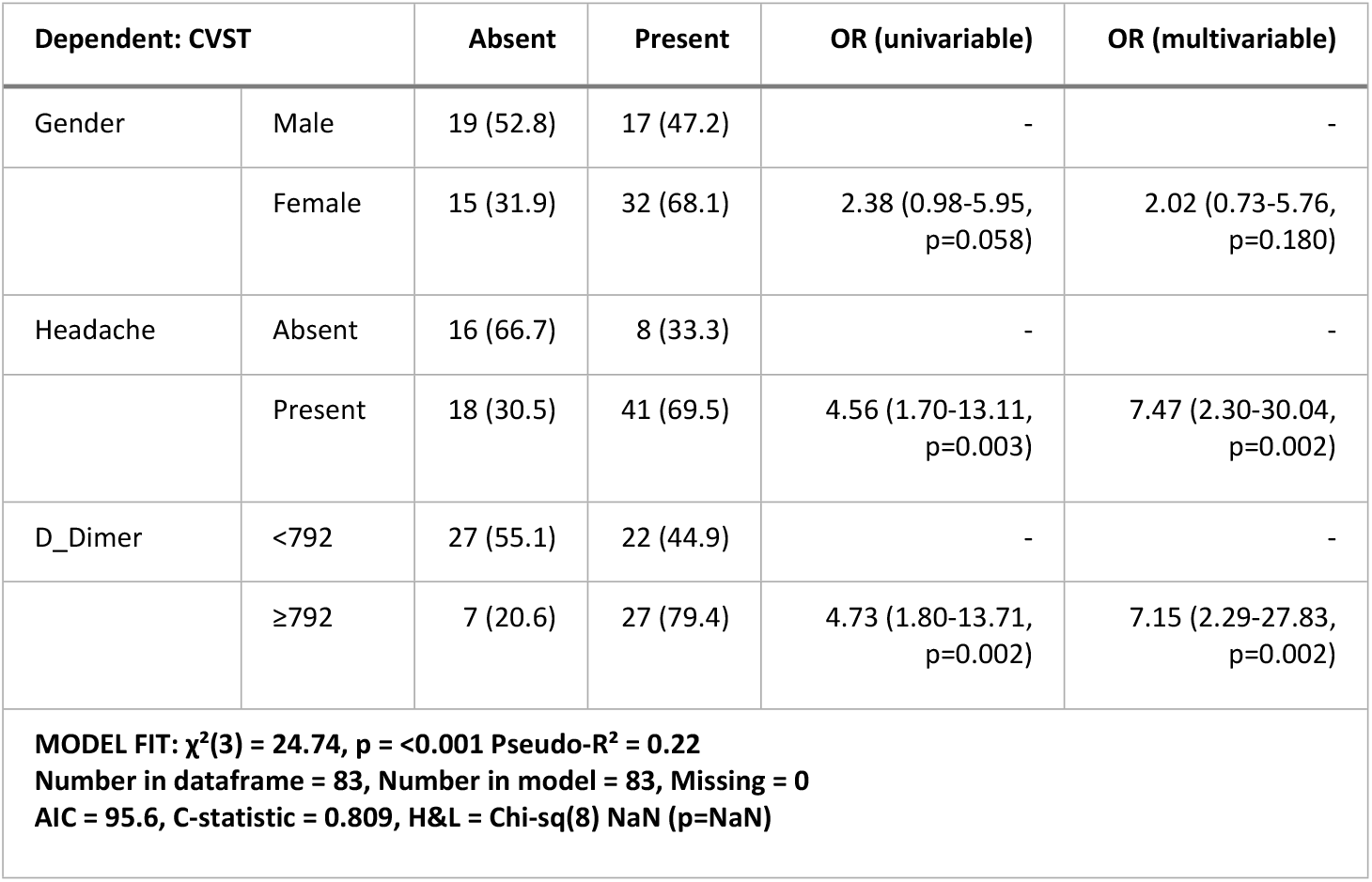
Regression analysis with all variables in the model.

| Dependent: CVST |  | Absent | Present | OR (univariable) | OR (multivariable) |
| --- | --- | --- | --- | --- | --- |
| Gender | Male | 19 (52.8) | 17 (47.2) | - | - |
|  | Female | 15 (31.9) | 32 (68.1) | 2.38 (0.98-5.95,<br>p=0.058) | 2.02 (0.73-5.76,<br>p=0.180) |
| Headache | Absent | 16 (66.7) | 8 (33.3) | - | - |
|  | Present | 18 (30.5) | 41 (69.5) | 4.56 (1.70-13.11,<br>p=0.003) | 7.47 (2.30-30.04,<br>p=0.002) |
| D_Dimer | <792 | 27 (55.1) | 22 (44.9) | - | - |
|  | ≥792 | 7 (20.6) | 27 (79.4) | 4.73 (1.80-13.71,<br>p=0.002) | 7.15 (2.29-27.83,<br>p=0.002) |
| **MODEL FIT: $\chi^2(3) = 24.74$, $p = <0.001$ Pseudo-$R^2 = 0.22$**<br>**Number in dataframe = 83, Number in model = 83, Missing = 0**<br>**AIC = 95.6, C-statistic = 0.809, H&L = Chi-sq(8) NaN (p=NaN)** |  |  |  |  |  |

If weightages are assigned to the individual factors by rounding off the odds ratio to the nearest integer, Female gender is assigned a score of 2, Headache (if present) is assigned a score of 7 and a D-Dimer level of ≥792 ng/mL is assigned a score of 7, with a maximum score possible being 16. (Table 3)

**Table 3:** Proposed score for the bedside prediction of CVST.

| Patient factor | Score Assigned |
| --- | --- |
| Female gender | 2 |
| Headache (if present) | 7 |
| D-Dimer level ≥ 792 ng/mL | 7 |

The total score for each participant was calculated and internally validated by ROC analysis (Fig 3), (Table 4). At a cutoff of Score ≥9, the proposed score has good diagnostic accuracy for CVST (AUROC = 0.809). The odds ratio (95% CI) for the presence of CVST when Score is ≥9 was 24.75 (3.13-195.81). The relative risk (95% CI) for the presence of CVST when Score is ≥9 was 2.08 (1.56-2.83).

**Table 4:**
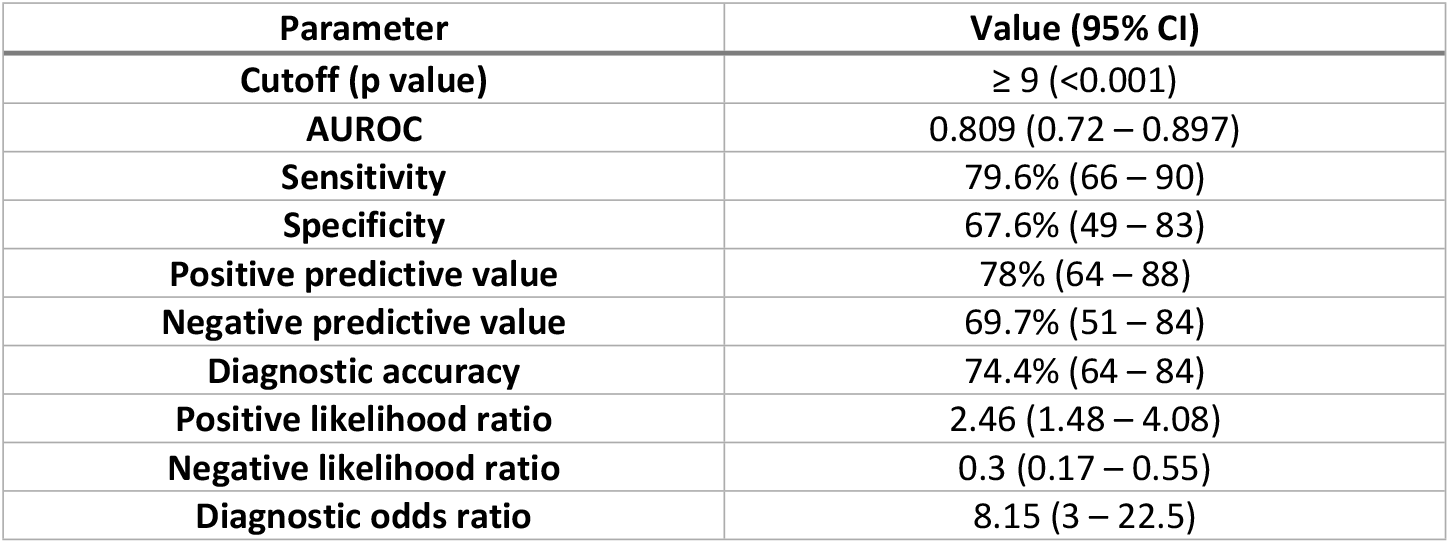
ROC curve analysis showing diagnostic performance of score in predicting CVST in suspected patients (n = 83)

**Fig 03:**
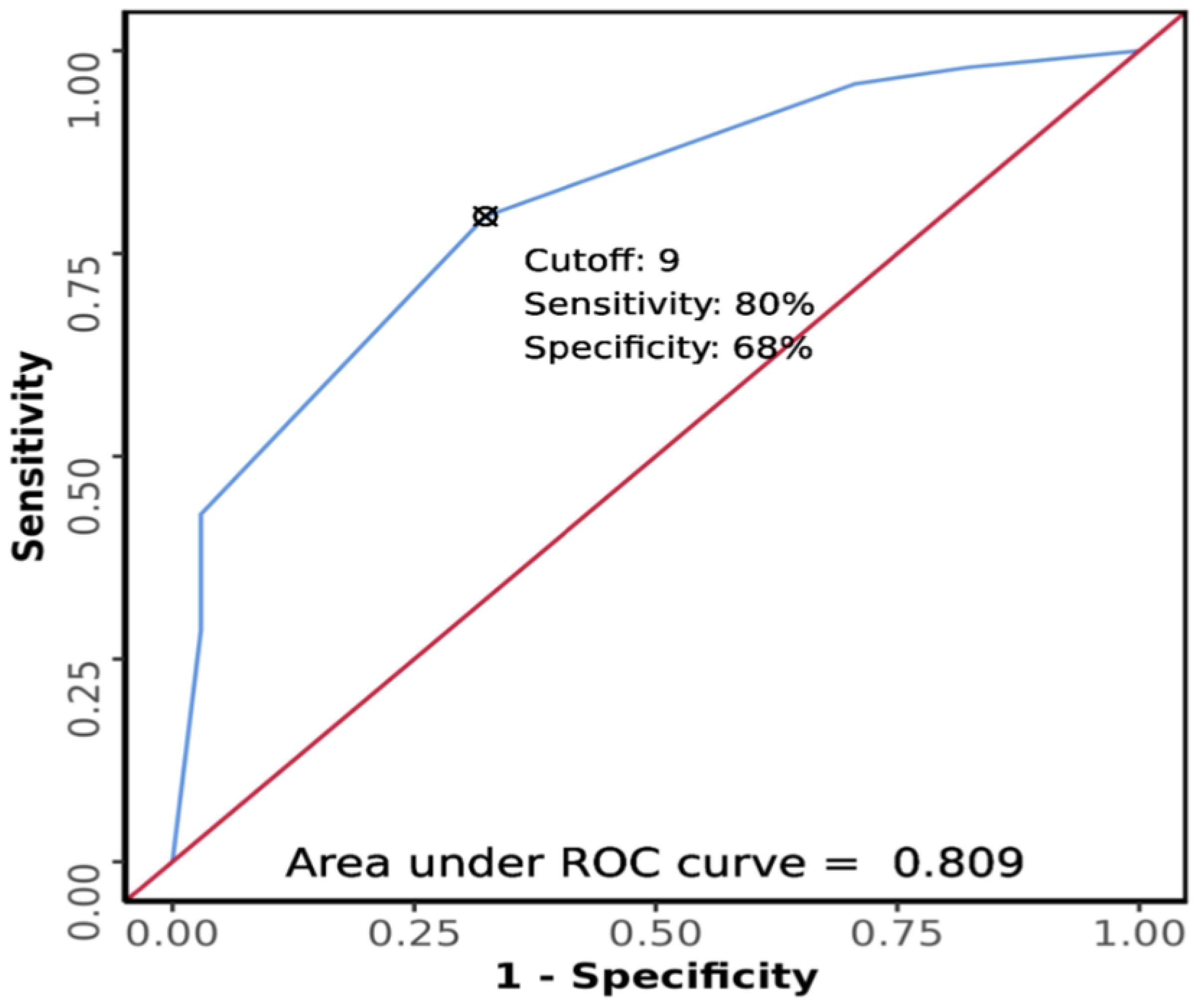
ROC curve analysis showing diagnostic performance of score in predicting CVST in suspected patients (n = 83)

## Discussion

A total of 83 patients suspected to be having CVST were enrolled. Forty-nine of the study participants had radiologically confirmed CVST, and 34 registered participants did not have CVST. The most common age group for CVST patients is 21 to 30 years (43%). India’s largest hospital-based cohort by Narayan et al. found a mean age of presentation is 31.3 years (8), and a recent study by J Kalita et al. in 2020, the median age was 28 years. (14) The landmark ISCVT cohort demonstrated that the median age for patients with CVST is 37 years. (12) All these studies highlight that the younger age group (21-40yrs) has a higher propensity for the development of CVST as in the present study.

Of all the patients who had CVST, two-third were females. The ISCVT cohort by Ferro et al. had nearly 75 % females in their study population (12) Though this is a known fact, the risk factors traditionally associated with CVST are seen in females, limited observational studies from India have shown an increased number of male patients with CVST. The study by Pai et al. had 57% of the study participants as males (15), while Narayan et al. had 53 % of their study population as males. (8) A recent study by Kalita et al. had 52% males in their study. (14) Most of these studies were hospital-based with a limited number of patients. Another reason could be a gender bias with a hospital seeking behaviour in males, as evident from the 2019-2020 census of a tertiary care centre in northern India. (16)

Though not statistically significant, the presence of Obesity (20%) and diabetes mellitus (14%) were more common in patients with CVST than those without CVST. Significant dehydration and fluid losses in the presence of hyperglycaemic emergencies such as diabetic ketoacidosis, hyperglycaemic non ketotic coma causes a relatively procoagulant state which may explain these findings. (17) The study by Pomp et al. in 2007 showed that obesity increases the risk of venous thromboembolism. (18) Obesity also increase the odds of a person developing CVST in the presence of an underlying procoagulant state. (13)

Approximately 83% of patients with CVST reported headache as a symptom on presentation. Narayan et al. reported headaches in 88 % of CVST patients (8), and Pai et al. had a headache in 62 % of theirs. (15) Nearly 89 % of patients had a headache in the study by Ferro et al. (12). Headache is multifactorial and is postulated to be due to stretching of walls of the Dural sinuses secondary to venous congestion. Hemorrhagic conversion of the lesion and the subsequent meningeal irritation and raised intracranial pressure may be a contributing factor. (1) In the Index study, 30% of enrolled CVST patients had papilledema, while 25 % had visual disturbance. The study by Ferro et al. had a similar percentage (30%)(12), while analysis by Pai et al. (15) and Narayan et al. (8) had 63 % and 64 % of CVST patients with papilledema, respectively. Papilledema in CVST is due to raised intracranial tension, and visual disturbances are a very late sign in papilledema as most of the damage to the retina in raised ICT occurs at the peripheral neurons, preserving the optic disk and macula until late in the course of illness. (20) The present study had 55% of CVST patients with seizure episodes on presentation, while other studies from India have fewer (30%,40%) of the CVST patients with this manifestation. (8), (15) Presence of seizures was commonly seen in postpartum CVST. (1), (12), (21). Motor deficits were more in the patients without CVST than those with CVST.(23% vs 20 %). This was probably due to the fact that more patients had an arterial stroke in the Non-CVST group, moreover, motor deficits that are readily discernible depend on the motor cortex being involved, which may not always be the case with CVST due to overlapping cerebral involvement involving multiple arterial territories. Hemiparesis noted in 50 % of CVST patients by Pai et al. (15), while Narayan et al.(8) and Ferro et al.(12) reported motor deficit in 28% and 37% CVST patients, respectively.

Patients with CVST, even those with known procoagulant states, were investigated for the presence of any new condition that could precipitate CVST. In the ISCVT cohort study of CVST patients, 13% did not have any identifiable risk factor. (12) In contrast, the present study found that 48% of CVST cases had no identifiable risk factor. Referral bias or lack of thorough workup due to the COVID pandemic may have contributed to higher proportion Idiopathic CVST cases in the present study. Postpartum state (16%) was the most common risk factor for CVST in this study, while Narayan et al.(8) and Pai et al.(15) identified 10% and 3 % of postpartum CVST cases, respectively. Other identifiable risk factors in this study was hyperhomocysteinemia (10%), followed by connective tissue disease (6%), OCP intake (4 %) and dehydration (4%). In a minority of patients, CNS infections, APLA, Protein S deficiency, malignancy, systemic Vasculitis were also identified. Two patients who were admitted with severe COVID 19 infection were enrolled in this study but then were later excluded as the disease duration was more than a week in these patients. None of the patients with CVST were COVID 19 vaccine recipients, the last patient enrolled in the study was at the time when the public vaccination programme was in its initial stages.

D Dimer is a fibrin degradation product, and though its presence in an elevated amount does not provide a host of information, low levels of D-Dimer aids in the exclusion of certain thrombotic disorders such as Deep vein thrombosis and pulmonary thromboembolism owing to its high negative predictive value. This idea was extrapolated and an attempt was made to evaluate the sensitivity of D-Dimer in exclusion of CVST patients in this study. (22) Role of D-Dimer in the diagnosis or exclusion of cerebral venous thrombosis is debatable, with the major limitation of the test being its non-specific nature, frequently leading to detection of elevated levels even in patients who did not have CVST. Most of the studies conducted in the past have demonstrated some utility of D-Dimer assay in exclusion of CVST in low risk patients who present with a headache, however, such a finding is often compounded by the fact that these patients will usually have another concomitant pathology which can contribute to the elevated D-Dimer levels. (23), (24), (25), (26) A Study by Ran Meng et al. in 2014 has shown the mean level of D Dimer was 968.9 ± 160 ng/mL in patients with CVST, which was significantly higher than in those without CVST. (27) A study analysing the utility of D Dimer in CVST patients by Tardy B et al. found that a value of <500 ng/mL was highly sensitive to rule out CVST. (26) Our study found that the median value of D Dimer was significantly elevated in the patients with CVST compared to those without CVST. (876 ng/mL vs 538 ng/mL) Further, on ROC analysis, D-dimer found to have low diagnostic accuracy in differentiating CVST from Non-CVST cases (AUROC 0.694).

For the computation of a clinical prediction rule, an initial univariate analysis was conducted that identified that Headache and an elevated D Dimer were significantly associated with the presence of CVST. Female gender was also considered for the above model as extensive studies by Leys D et al. (11), Ferro et al. (12) has clearly shown a significantly higher association of CVST with the female gender. The odds ratio for each of these three variables were computed using Multivariate regression analysis, which were rounded off to the nearest integer and assigned as a ‘weightage’ (headache 7 points, Female gender 2 points, D Dimer >792 ng/mL 7 points). This scoring was internally validated in the study population, and ROC analysis identified the cut-off score of ≥9, above which the diagnosis of CVST was highly likely (AUROC = 0.809). This methodology of generating the clinical prediction model is analogous to the 2020 study by Heldner et al., which demonstrated that the addition of D Dimer as a component in the clinical scoring improved the overall diagnostic accuracy of the score (AUROC 0.889 without D Dimer to AUROC 0.937 with the inclusion of D Dimer). (29)

## Conclusions

1. ROC analysis revealed that D-Dimer assay had a poor diagnostic accuracy in differentiating CVST from non-CVST patients (AUROC = 0.694), however, a D-Dimer level of <300 ng/mL had a sensitivity of 90% to exclude the diagnosis of CVST.
2. After logistic regression analysis, a clinical decision rule with a total score of 16 and individual components of Female gender (2 points), Presence of headache (7 points), presence of D-Dimer levels of ≥ 792 ng/mL (7 points) was devised, a score of ≥ 9 had good diagnostic accuracy for the prediction of CVST. (AUROC = 0.809)

### Limitations of the study

1. The study was conducted at a single centre, since referral bias may be a factor in such a centre, it may not be representative of the entire population. Multicentre studies would be ideal for a study with such a goal.
2. The sample size was limited to 83 patients, inclusion of a larger number of participants in future studies is needed for the information obtained to be generalizable.
3. The study period was for 17 months and follow up of the enrolled participants could not be carried out
4. Patient’s genetic factors as well as the presence of infections, inflammatory response of the body, co-administered drugs can interfere with the measurement of quantitative D-Dimer assay, though any clinically possible interference with the results was thought of, given the logistical and resource constrains, it is difficult to exclude all the factors that may lead to altered levels of D-Dimer.
5. In the computation of clinical decision rule, the presence of Female gender was taken as a component of the decision rule, in our study, however, the presence of female gender was not significantly associated with the presence of CVST with the p value just approaching the cut off of 0.05. (actual p = 0.055)
6. The proposed clinical decision rule is internally validated in the study and needs to be externally validated in subsequent studies to standardise the findings.

## Data Availability

All relevant data are within the manuscript.

## Acknowledgements

The authors would like to acknowledge the help provided by the coagulation laboratory staff, Mr. Chanderhans and Ms. Anita Kler. We would also like to extend our gratitude to Junior administrative assistant, Mr. Ajay Kumar for his input in statistical analysis.

## References

1. Dash D, Prasad K, Joseph L. Cerebral venous thrombosis: An Indian perspective. Neurol India. 2015;63(3):318–28.

2. Ferro JM, Canhão P. Cerebral venous sinus thrombosis: update on diagnosis and management. Curr Cardiol Rep. 2014;16(9):523.

3. Bansal BC, Gupta RR, Prakash C. Stroke during pregnancy and puerperium in young females below the age of 40 years as a result of cerebral venous/venous sinus thrombosis. Jpn Heart J 1980;21:171–73

4. Banerjee AK, Varma M, Vasista RK, Chopra JS. Cerebrovascular disease in north-west India: A study of necropsy material. J Neurol Neurosurg Psychiatry 1989;52:512–5

5. Stam J. Thrombosis of the cerebral veins and sinuses. N Engl J Med 2005; 352: 1791–8

6. Chiewvit P, Piyapittayanan S, Poungvarin N. Cerebral venous thrombosis: diagnosis dilemma. Neurol Int. 2011;3(3):13

7. Cucchiara B, Messe S, Taylor R, Clarke J, Pollak E. Utility of D-dimer in the diagnosis of cerebral venous sinus thrombosis. J Thromb Haemost 2005; 3: 387–9

8. Narayan D, Kaul S, Ravishankar K, Suryaprabha T, Bandaru VCSS, Mridula KR, et al. Risk factors, clinical profile, and long-term outcome of 428 patients of cerebral sinus venous thrombosis. Neurol India. 2012 Mar-Apr;60(2):154–9

9. Linkins L, Lapner ST. Review of D-dimer testing: Good, Bad, and Ugly. Int J Lab Hematol. 2017 May;39 Suppl 1:98–103

10. Fenn Buderer NM. Statistical methodology: incorporating the prevalence of disease into the sample size calculation for sensitivity and specificity. Acad Emerg Med. 1996;3(9):895–900.

11. Leys D, Cordonnier C. Cerebral venous thrombosis: Update on clinical manifestations, diagnosis and management. Ann Indian Acad Neurol. 2008;11.

12. Ferro JM, Canhão P, Stam J, Bousser MG, Barinagarrementeria F. Prognosis of Cerebral Vein and Dural Sinus Thrombosis: Results of the International Study on Cerebral Vein and Dural Sinus Thrombosis (ISCVT). Stroke. 2004;35(3):664–70.

13. Zuurbier SM, Arnold M, Middeldorp S, Broeg-Morvay A, Silvis SM, Heldner MR, et al. Risk of Cerebral Venous Thrombosis in Obese Women. JAMA Neurol. 2016 May 1;73(5):579–84

14. Kalita J, Singh VK, Misra UK. A study of hyperhomocysteinemia in cerebral venous thrombosis. Indian J Med Res 2020; 152:584–94.

15. Pai N, Ghosh K, Shetty S. Hereditary thrombophilia in cerebral venous thrombosis: A study from India. Blood Coagul Fibrinolysis. 2013;24(5):540–3.

16. Sahni D et al. 2020. 53^rd^ Annual Report, PGIMER.

17. Keane S, Gallagher A, Ackroyd S, et al Cerebral venous thrombosis during diabetic ketoacidosis Archives of Disease in Childhood 2002;86:204–205

18. Pomp ER, le Cessie S, Rosendaal FR, Doggen CJ. Risk of venous thrombosis: obesity and its joint effect with oral contraceptive use and prothrombotic mutations. Br J Haematol. 2007;139 (2):289–29

19. Ferro JM, Aguiar de Sousa D. Cerebral Venous Thrombosis: an Update. Curr Neurol Neurosci Rep. 2019 Aug 23;19(10):74.

20. Wall M, George D. Visual Loss in Pseudotumor Cerebri: Incidence and Defects Related to Visual Field Strategy. Arch Neurol. 1987;44(2):170–175 a. Ferro JM, Bousser MG, Canhão P, Coutinho JM, Crassard I, Dentali F, et al. European Stroke Organization guideline for the diagnosis and treatment of cerebral venous thrombosis Endorsed by the European Academy of Neurology. Eur Stroke J. 2017;2(3):195–221.

21. Couturaud F, Kearon C, Bates SM, Ginsberg JS. Decrease in sensitivity of D-dimer for acute venous thromboembolism after starting anticoagulant therapy. Blood Coagul Fibrinolysis. 2002 Apr;13(3):241–6

22. Caplan LR. Caplan’s Stroke. 5th edition. United Kingdom: Cambridge University Press; 2016

23. Fernandes CJ, Morinaga LTK, Alves JL Jr, Castro MA, Calderaro D, Jardim CVP, Souza R. Cancer-associated thrombosis: the when, how and why. Eur Respir Rev. 2019 Mar 27;28(151):180119

24. Alons IME, Jellema K, Wermer MJH, Algra A. D DIMER for the exclusion of cerebral venous thrombosis: A meta-analysis of low risk patients with isolated headache. BMC Neurol. 2015;15(1):1–7.

25. Smith, E., & Kumar, V. (2018). BET 1: Does a normal D-dimer rule out cerebral venous sinus thrombosis (CVST). Emergency Medicine Journal. 2018

26. Meng R, Wang X, Hussain M, Dornbos D 3rd, Meng L, Liu Y, Wu Y, Ning M, Ferdinando S B, Lo EH, Ding Y, Ji X. Evaluation of plasma D-dimer plus fibrinogen in predicting acute CVST. Int J Stroke. 2014 Feb;9(2):166–73.

27. Tardy B, et al. D-Dimer levels in patients with suspected acute cerebral venous thrombosis. Am j Med. 2002;113:238–241

28. Heldner MR, Zuurbier SM, Li B, Von Martial R, Meijers JCM, Zimmermann R, Volbers B, Jung S, El-Koussy M, Fischer U, Kohler HP, Schroeder V, Coutinho JM, Arnold M. Prediction of cerebral venous thrombosis with a new clinical score and D-dimer levels. Neurology. 2020 Aug 18;95(7):e898–e909

